# Development and Validation of a Sleep Disorder Risk Prediction Model in Young and Middle-Aged Adults With Anemia

**DOI:** 10.64898/2026.10.12.26365352

**Authors:** Yan Liu, Cangyuan Zhang

**Affiliations:** Shandong Public Health Clinical Center, Shandong University, Jinan, Shandong 250013, China

**Keywords:** Anemia, Sleep disorder, NHANES database, Nomogram

## Abstract

**Objective:** This study aimed to develop and validate a predictive model for sleep disorders in young and middle-aged adults with anemia, providing evidence for early identification, risk stratification, and individualized sleep management.

**Methods:** Data in the NHANES were analyzed and randomly divided into development and validation cohorts. LASSO regression was used to screen optimal predictors in the development cohort, followed by univariate and multivariate logistic regression to identify independent predictors and construct a nomogram. Model performance was evaluated using receiver operating characteristic curves, calibration curves, Brier scores, and decision curve analysis.

**Results:** The weighted prevalence of sleep disorders among young and middle-aged adults with anemia was 10.19%. Multivariate logistic regression identified seven independent predictors: sex, monocyte count, asthma, overweight, arthritis, congestive heart failure, and hypertension. The model showed good discrimination, with area under the curve values of 0.817 in the development cohort and 0.791 in the validation cohort. Calibration was satisfactory, with Brier scores of 0.074 and 0.075, respectively. Decision curve analysis indicated favorable clinical net benefit in both cohorts.

**Conclusion:** The proposed predictive model demonstrated good discrimination, calibration, and may help healthcare providers identify young and middle-aged adults with anemia who are at high risk for sleep disorders.

## 1 Introduction

Anemia is one of the most prevalent hematological disorders on a global scale, constituting a substantial public health concern. The 2021 Global Burden of Disease Study revealed that the global prevalence of anemia is approximately 24.3%, affecting approximately 1.92 billion people. The disease burden varies significantly across different age groups, genders, and regions [1, 2]. In accordance with the criteria established by the World Health Organization (WHO), anemia is generally characterized as a condition in which the hemoglobin (Hb) level falls below the standard range [3]. Specifically, the Hb level is defined as less than 12.0 g/dL in adult female patients and less than 13.0 g/dL in adult male patients. The etiology of anemia is complex, and its clinical manifestations depend on the type, severity and rate of onset of the anemia. The symptoms experienced by patients with this condition are diverse and include fatigue, weakness, shortness of breath, decreased concentration, reduced exercise tolerance, and limitations in activities of daily living [4–6].

The quality of sleep is influenced by a variety of individual and environmental factors. Sleep disorders are not a single disease but rather a group of clinical conditions characterized by abnormalities in sleep quantity, quality, rhythm, or structure [7, 8]. A plethora of studies have demonstrated an association between a short sleep duration and a multitude of adverse health outcomes, including cardiovascular events, metabolic abnormalities, and an elevated risk of all-cause mortality [9,10]. Consequently, sleep disorders have the potential to impact not only the subjective experience of rest but also serve as a significant phenotype for the development of chronic diseases and the deterioration of health status.

The potential for a bidirectional relationship between anemia and sleep disorders is a salient consideration. Tissue hypoxia and fatigue caused by anemia have been demonstrated to exacerbate nocturnal discomfort and impair daytime functioning [11,12]. In turn, chronic sleep deprivation has been shown to amplify fatigue and the burden of chronic diseases through immune-inflammatory activation, sympathetic nervous system excitation, and endocrine disruption [11,12]. A cross-sectional study and meta-analysis by Neumann et al. demonstrated a significant association between anemia in adults and an increased incidence of insomnia [13].

Adults in the early and mid-adult years find themselves in a phase of life typified by the convergence of significant academic, professional and familial obligations. A number of factors have been identified as potential risk factors for the development of sleep disorders. These include sleep deprivation, psychological stress, sedentary behaviour, obesity, and the early onset of chronic diseases [14, 15]. At present, systematic reviews on risk factors for sleep disorders among young and middle-aged individuals with anemia remain limited, and there is a paucity of predictive tools for individualized risk assessment. Consequently, the development and validation of a risk prediction model for sleep disorders in this population will facilitate the early identification of high-risk patients and establish a foundation for subsequent personalized interventions and public health management.

## 2 Methods

### 2.1 Data sources and study population

The data for this study were obtained from the National Health and Nutrition Examination Survey (NHANES) website. NHANES is a comprehensive, nationally representative survey designed to assess and understand the health, well-being, and nutritional status of individuals residing in the United States. The collection of data is achieved through a variety of methods, including interviews, physical examinations and laboratory tests. The objective of this process is to provide a comprehensive understanding of the health and nutritional characteristics of the population. The survey is available online at https://wwwn.cdc.gov/nchs/nhanes/.

The NHANES study protocol has been approved by the NCHS Institutional Review Board, and all participants signed written informed consent prior to the survey. Relevant ethical approval and data access procedures can be found on the official NHANES website.

This study included participants aged 20–60 years from the 2005–2018 NHANES cohorts and further screened for those meeting the diagnostic criteria for anemia (see Figure 1). Anemia is defined according to the relevant standards of WHO [3]: male hemoglobin level<13.0 g/dL, female hemoglobin level<12.0 g/dL. Exclusion criteria included: (1) lack of information related to hemoglobin or sleep disorders; (2) Incomplete data of key predictive variables; (3) Missing sampling weights, stratified or clustered variables; (4) Other data records that do not meet the requirements of research and analysis. Finally, young and middle-aged patients with anemia who met the criteria were included for follow-up analysis.

**Figure 1.**
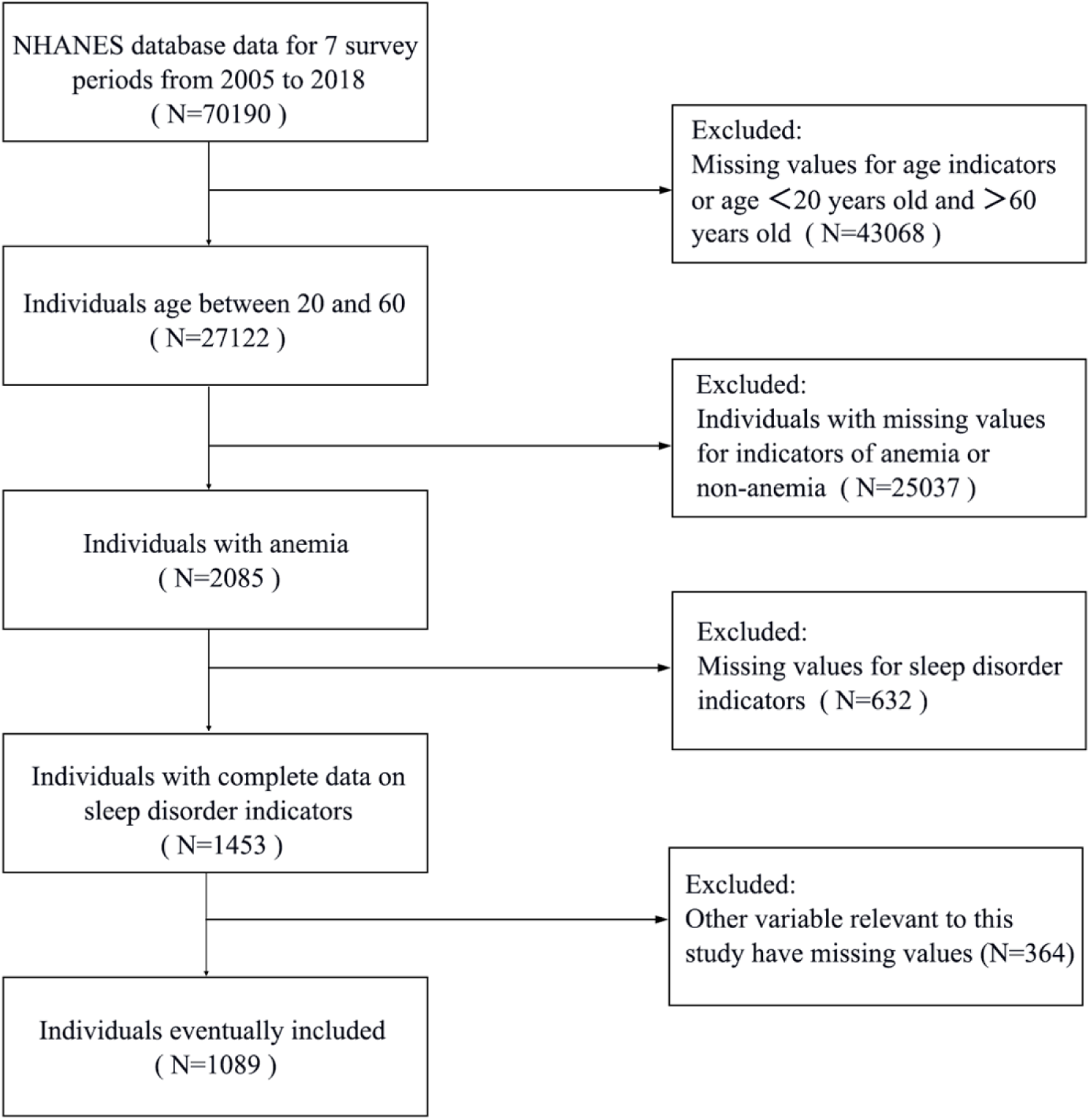
Research object screening flow chart.

### 2.2 Definition and interpretation of indicators

The candidate predictor variables encompassed in this study comprised demographic characteristics, anthropometric measurements, laboratory test results, and questionnaire-based variables.

The demographic variables encompassed age, sex, race/ethnicity, educational attainment, marital status, and poverty income ratio (PIR). The categorization of race/ethnicity encompassed Mexican American, other Hispanic, non-Hispanic White, non-Hispanic Black, and other races. Educational attainment was categorized as follows: below ninth grade, ninth–eleventh grade, high school graduate or General Educational Development (GED) certificate, some college or associate’s degree, and college graduate or higher. The marital status of the participants was categorised as follows: married/cohabiting, widowed/divorced/separated, and unmarried. The utilisation of PIR (Poverty Index Rating) was employed as a metric to reflect household economic status. The physical measurements encompassed the body mass index (BMI).

Laboratory test measures encompass a range of parameters, including white blood cell count, red blood cell count, hemoglobin level, platelet count, lymphocyte count, monocyte count, albumin level, uric acid level, lactate dehydrogenase (LDH) level, and direct high-density lipoprotein cholesterol (HDL-C) level.

The questionnaire under review included a series of variables pertaining to various health concerns, including asthma, arthritis, congestive heart failure, stroke, hypertension, diabetes, smoking status, and urinary incontinence. The definitions of these variables were derived from the participants’ self-reported history of physician diagnoses or related health behaviors as indicated in the NHANES questionnaire. The categorization of these dichotomous variables was conducted employing a binary coding system, with the assigned codes “No” and “Yes” employed uniformly.

Sleep disturbance was the primary outcome variable in this study and was operationally defined based on self-reported information regarding sleep difficulties in the NHANES sleep questionnaire [16]. Participants who reported experiencing sleep difficulties or having been diagnosed with sleep-related conditions were categorized into the sleep disturbance group. Conversely, those who did not report sleep disturbances or have a documented sleep-related condition were classified into the non-sleep disturbance group.

### 2.3 Statistical analysis

All statistical analyses were performed using R software (version 4.4.1). In order to accurately reflect the overall health status of the U.S. population, the sample statistics were weighted in order to address potential under- or over-representation of different subgroups during the sampling process. The “WTMEC2YR” function adjusts the data to reflect population weights, thereby enhancing the accuracy and reliability of the statistical analysis.

Weighted continuous variables are presented as means and standard errors (SE), and between-group differences were compared using independent samples t-tests; categorical variables are presented as percentages and standard errors, and between-group comparisons were performed using Pearson’s χ² test. Initially, the candidate variables were subjected to screening utilizing the LASSO regression. Subsequently, weighted univariate and multivariate logistic regression analyses were conducted to identify independent risk factors for sleep disorders in young and middle-aged patients with anemia, and a risk plot for sleep disorders was constructed [14]. The evaluation of model discrimination was conducted by utilizing the ROC curve and the area under the curve. Calibration was assessed employing the Brier score and Spiegelhalter’s z-test. The validation of clinical utility was achieved through DCA. All statistical tests were two-sided, and a *P*-value of less than 0.05 was considered to be statistically significant.

## 3 Results

### 3.1 Comparison of data between development group and validation group

The weighted average age of the 1,089 respondents was 40.20 years, with a standard error of 0.43 years. The gender distribution of the population was as follows: 18.38% male and 81.62% female. With respect to educational attainment, the weighted proportions were as follows: below high school (18.93%), high school graduate/GED or equivalent (20.10%), some college or associate’s degree (35.45%), and bachelor’s degree or higher (25.52%). As demonstrated in Table 1, a comprehensive overview of the weighted data is provided. The weighted prevalence of sleep disorders among the 1,089 participants was 10.19%. The 1,089 patients were randomly assigned to a development group (n = 763) and a validation group (n = 326) in a 7:3 ratio. Table 1 presents the baseline characteristics of patients in the training and validation groups. The findings demonstrated that there were no statistically significant differences between the two groups (*P* > 0.05), thus indicating that the random assignment of the dataset was appropriate and suitable for subsequent analysis.

**Table 1.**
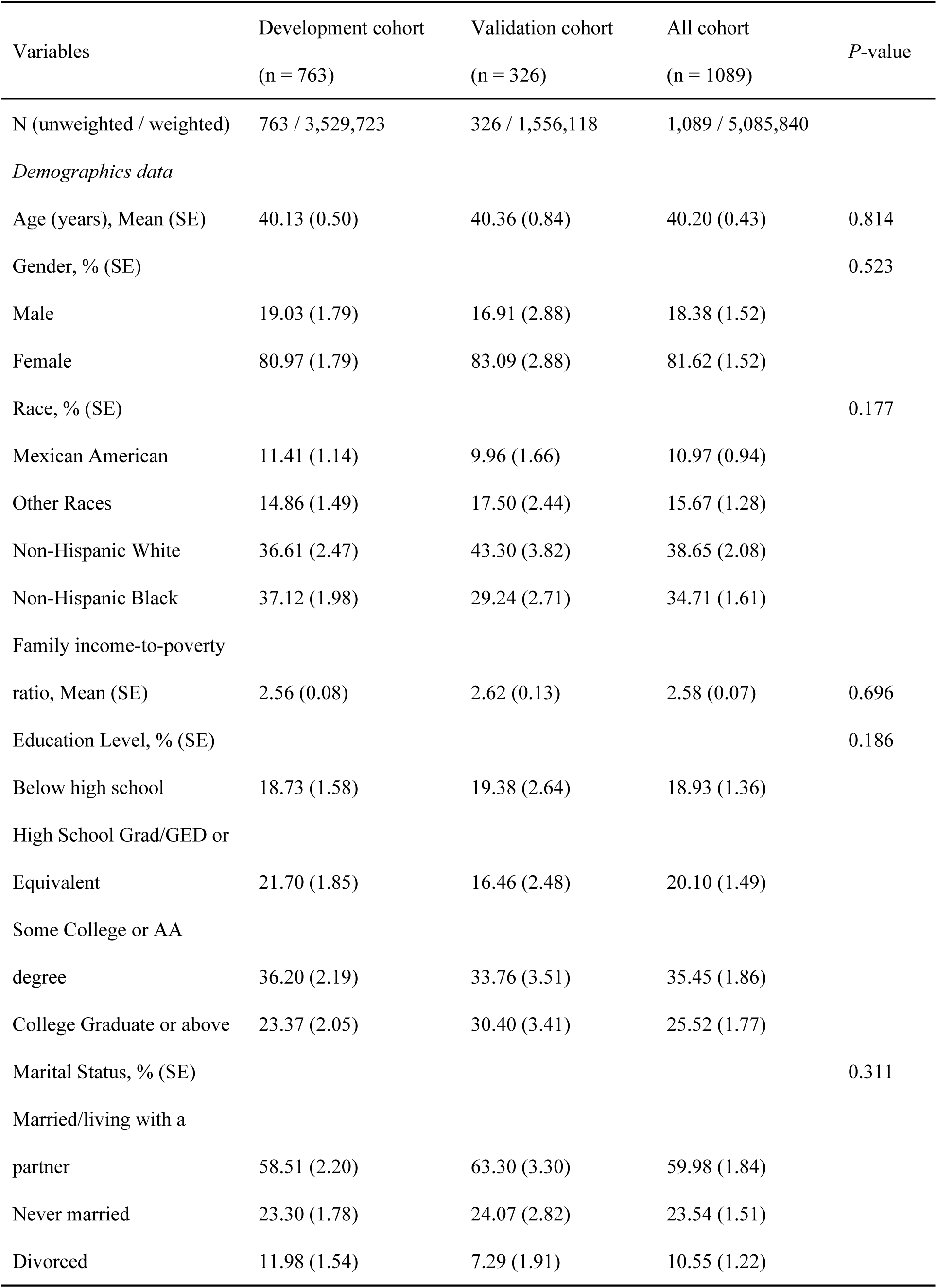
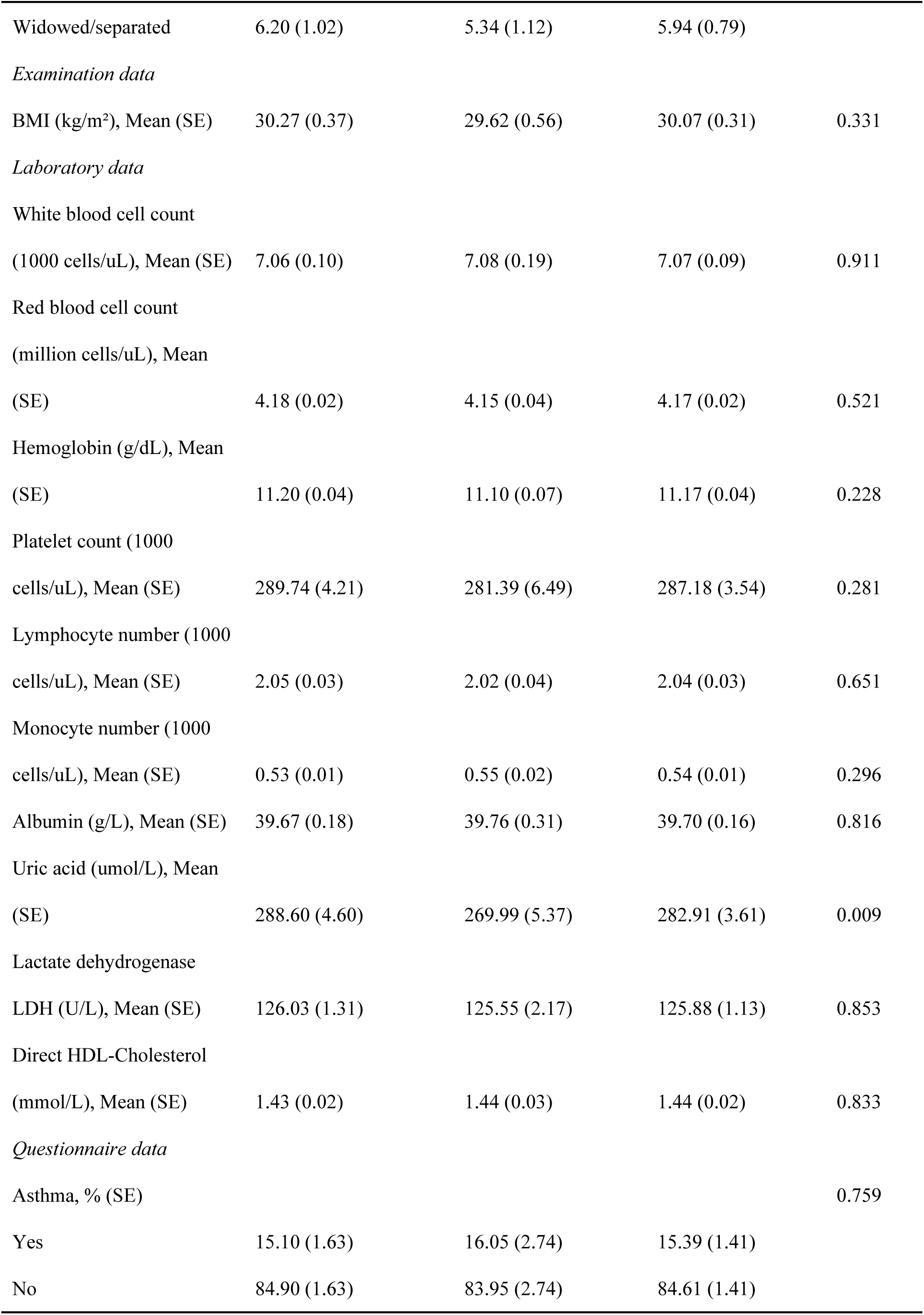
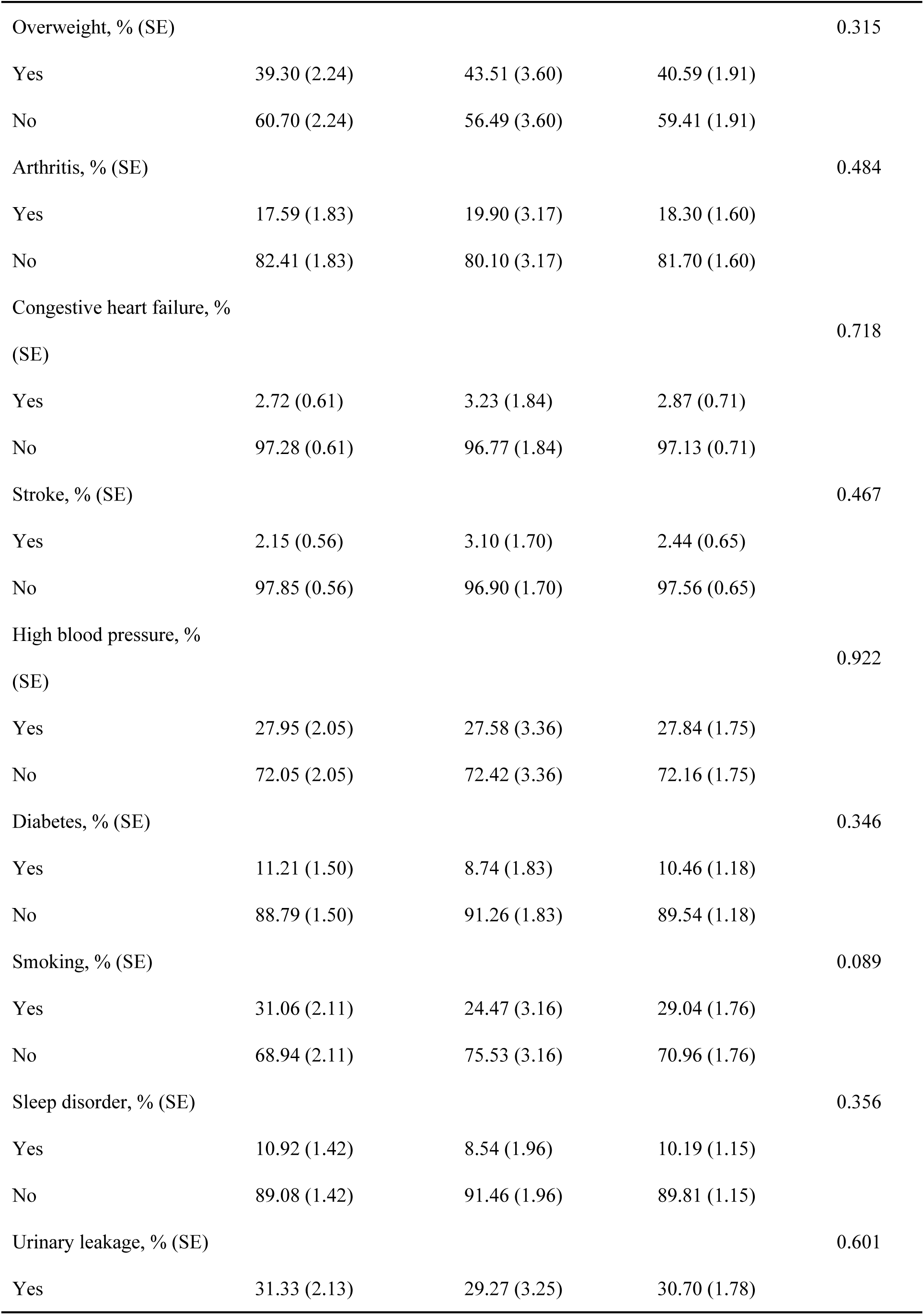
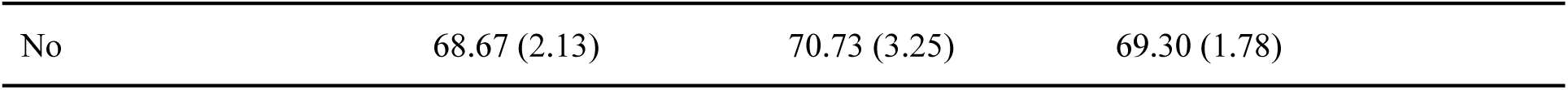
Comparison of data between development cohort and validation cohort.

| Variables | Development cohort<br>(n = 763) | Validation cohort<br>(n = 326) | All cohort<br>(n = 1089) | P-value |
| --- | --- | --- | --- | --- |
| N (unweighted / weighted) | 763 / 3,529,723 | 326 / 1,556,118 | 1,089 / 5,085,840 |  |
| *Demographics data* |  |  |  |  |
| Age (years), Mean (SE) | 40.13 (0.50) | 40.36 (0.84) | 40.20 (0.43) | 0.814 |
| Gender, % (SE) |  |  |  | 0.523 |
| Male | 19.03 (1.79) | 16.91 (2.88) | 18.38 (1.52) |  |
| Female | 80.97 (1.79) | 83.09 (2.88) | 81.62 (1.52) |  |
| Race, % (SE) |  |  |  | 0.177 |
| Mexican American | 11.41 (1.14) | 9.96 (1.66) | 10.97 (0.94) |  |
| Other Races | 14.86 (1.49) | 17.50 (2.44) | 15.67 (1.28) |  |
| Non-Hispanic White | 36.61 (2.47) | 43.30 (3.82) | 38.65 (2.08) |  |
| Non-Hispanic Black | 37.12 (1.98) | 29.24 (2.71) | 34.71 (1.61) |  |
| Family income-to-poverty<br>ratio, Mean (SE) | 2.56 (0.08) | 2.62 (0.13) | 2.58 (0.07) | 0.696 |
| Education Level, % (SE) |  |  |  | 0.186 |
| Below high school | 18.73 (1.58) | 19.38 (2.64) | 18.93 (1.36) |  |
| High School Grad/GED or<br>Equivalent | 21.70 (1.85) | 16.46 (2.48) | 20.10 (1.49) |  |
| Some College or AA<br>degree | 36.20 (2.19) | 33.76 (3.51) | 35.45 (1.86) |  |
| College Graduate or above | 23.37 (2.05) | 30.40 (3.41) | 25.52 (1.77) |  |
| Marital Status, % (SE) |  |  |  | 0.311 |
| Married/living with a<br>partner | 58.51 (2.20) | 63.30 (3.30) | 59.98 (1.84) |  |
| Never married | 23.30 (1.78) | 24.07 (2.82) | 23.54 (1.51) |  |
| Divorced | 11.98 (1.54) | 7.29 (1.91) | 10.55 (1.22) |  |
| Widowed/separated | 6.20 (1.02) | 5.34 (1.12) | 5.94 (0.79) |  |
| *Examination data* |  |  |  |  |
| BMI (kg/m<sup>2</sup>), Mean (SE) | 30.27 (0.37) | 29.62 (0.56) | 30.07 (0.31) | 0.331 |
| *Laboratory data* |  |  |  |  |
| White blood cell count |  |  |  |  |
| (1000 cells/uL), Mean (SE) | 7.06 (0.10) | 7.08 (0.19) | 7.07 (0.09) | 0.911 |
| Red blood cell count |  |  |  |  |
| (million cells/uL), Mean |  |  |  |  |
| (SE) | 4.18 (0.02) | 4.15 (0.04) | 4.17 (0.02) | 0.521 |
| Hemoglobin (g/dL), Mean |  |  |  |  |
| (SE) | 11.20 (0.04) | 11.10 (0.07) | 11.17 (0.04) | 0.228 |
| Platelet count (1000 |  |  |  |  |
| cells/uL), Mean (SE) | 289.74 (4.21) | 281.39 (6.49) | 287.18 (3.54) | 0.281 |
| Lymphocyte number (1000 |  |  |  |  |
| cells/uL), Mean (SE) | 2.05 (0.03) | 2.02 (0.04) | 2.04 (0.03) | 0.651 |
| Monocyte number (1000 |  |  |  |  |
| cells/uL), Mean (SE) | 0.53 (0.01) | 0.55 (0.02) | 0.54 (0.01) | 0.296 |
| Albumin (g/L), Mean (SE) | 39.67 (0.18) | 39.76 (0.31) | 39.70 (0.16) | 0.816 |
| Uric acid (umol/L), Mean |  |  |  |  |
| (SE) | 288.60 (4.60) | 269.99 (5.37) | 282.91 (3.61) | 0.009 |
| Lactate dehydrogenase |  |  |  |  |
| LDH (U/L), Mean (SE) | 126.03 (1.31) | 125.55 (2.17) | 125.88 (1.13) | 0.853 |
| Direct HDL-Cholesterol |  |  |  |  |
| (mmol/L), Mean (SE) | 1.43 (0.02) | 1.44 (0.03) | 1.44 (0.02) | 0.833 |
| *Questionnaire data* |  |  |  |  |
| Asthma, % (SE) |  |  |  | 0.759 |
| Yes | 15.10 (1.63) | 16.05 (2.74) | 15.39 (1.41) |  |
| No | 84.90 (1.63) | 83.95 (2.74) | 84.61 (1.41) |  |
| Overweight, % (SE) |  |  |  | 0.315 |
| Yes | 39.30 (2.24) | 43.51 (3.60) | 40.59 (1.91) |  |
| No | 60.70 (2.24) | 56.49 (3.60) | 59.41 (1.91) |  |
| Arthritis, % (SE) |  |  |  | 0.484 |
| Yes | 17.59 (1.83) | 19.90 (3.17) | 18.30 (1.60) |  |
| No | 82.41 (1.83) | 80.10 (3.17) | 81.70 (1.60) |  |
| Congestive heart failure, %<br>(SE) |  |  |  | 0.718 |
| Yes | 2.72 (0.61) | 3.23 (1.84) | 2.87 (0.71) |  |
| No | 97.28 (0.61) | 96.77 (1.84) | 97.13 (0.71) |  |
| Stroke, % (SE) |  |  |  | 0.467 |
| Yes | 2.15 (0.56) | 3.10 (1.70) | 2.44 (0.65) |  |
| No | 97.85 (0.56) | 96.90 (1.70) | 97.56 (0.65) |  |
| High blood pressure, %<br>(SE) |  |  |  | 0.922 |
| Yes | 27.95 (2.05) | 27.58 (3.36) | 27.84 (1.75) |  |
| No | 72.05 (2.05) | 72.42 (3.36) | 72.16 (1.75) |  |
| Diabetes, % (SE) |  |  |  | 0.346 |
| Yes | 11.21 (1.50) | 8.74 (1.83) | 10.46 (1.18) |  |
| No | 88.79 (1.50) | 91.26 (1.83) | 89.54 (1.18) |  |
| Smoking, % (SE) |  |  |  | 0.089 |
| Yes | 31.06 (2.11) | 24.47 (3.16) | 29.04 (1.76) |  |
| No | 68.94 (2.11) | 75.53 (3.16) | 70.96 (1.76) |  |
| Sleep disorder, % (SE) |  |  |  | 0.356 |
| Yes | 10.92 (1.42) | 8.54 (1.96) | 10.19 (1.15) |  |
| No | 89.08 (1.42) | 91.46 (1.96) | 89.81 (1.15) |  |
| Urinary leakage, % (SE) |  |  |  | 0.601 |
| Yes | 31.33 (2.13) | 29.27 (3.25) | 30.70 (1.78) |  |

|  |  |  |  |
| --- | --- | --- | --- |
| No | 68.67 (2.13) | 70.73 (3.25) | 69.30 (1.78) |

### 3.2 Selection of predictors and nomogram construction

LASSO regression analysis was employed to identify the optimal predictors (see Figure 2). These predictors were then used in univariate and multivariate logistic regression analyses. The results indicated that gender (*P* = 0.027), monocyte count (*P* = 0.009), asthma (*P* = 0.003), overweight (*P* < 0.001), arthritis (*P* = 0.009), congestive heart failure (*P* = 0.033), and hypertension (*P* = 0.012) were identified as independent risk factors for sleep disorders (see Table 2 and Table 3).

**Figure 2.**
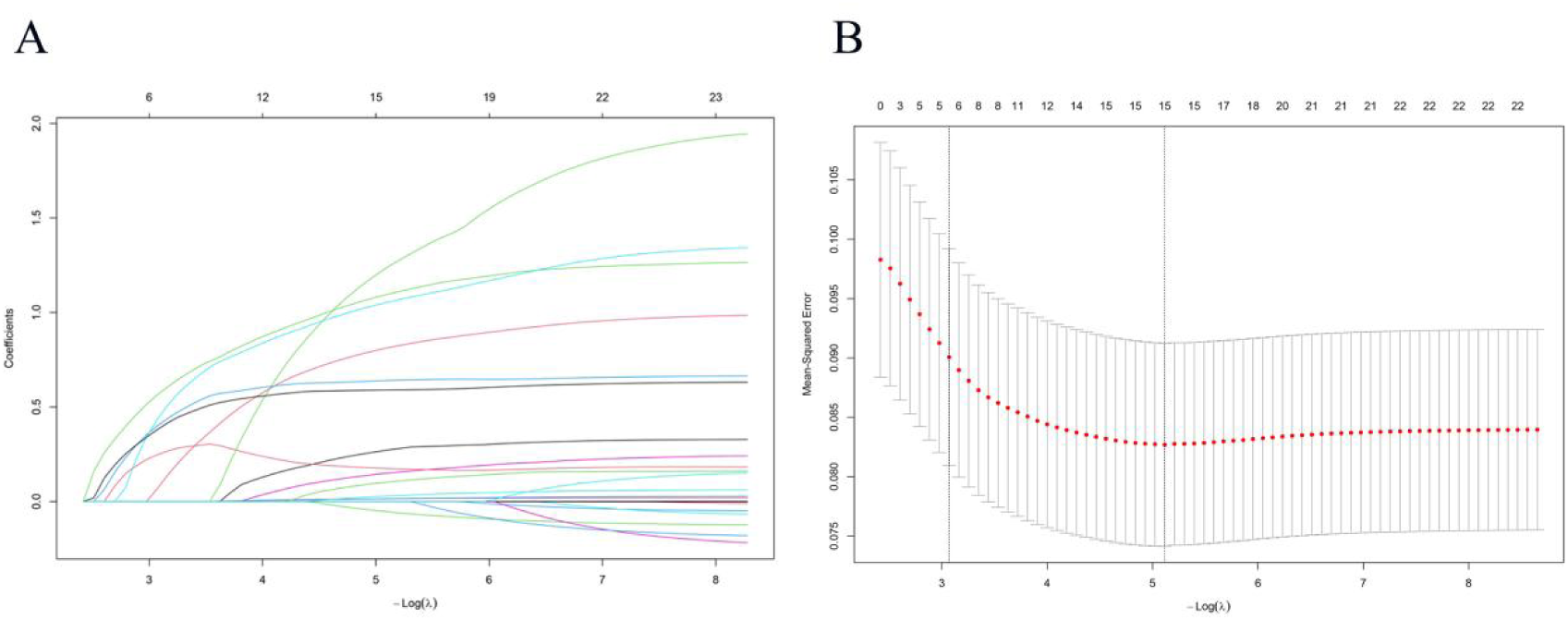
Selection of potential predictors through the LASSO regression model.

**Table 2.** Results of weighted univariate Logistic regression analysis of sleep disorder in young and middle-aged patients in the development cohort.

| Variables | Estimate | Std. Error | Z-value | P-value | OR | CI (2.5%) | CI (97.5%) |
| --- | --- | --- | --- | --- | --- | --- | --- |
| *Demographics data* |  |  |  |  |  |  |  |
| Age (years) | 0.062 | 0.012 | 5.326 | 0.001* | 1.063 | 1.040 | 1.089 |
| Gender |  |  |  |  |  |  |  |
| Male | Ref |  |  |  |  |  |  |
| Female | -0.909 | 0.251 | -3.625 | 0.001* | 0.403 | 0.248 | 0.665 |
| Race |  |  |  |  |  |  |  |
| Mexican American | Ref |  |  |  |  |  |  |
| Other Races | 0.240 | 0.483 | 0.496 | 0.620 | 1.271 | 0.496 | 3.393 |
| Non-Hispanic White | 0.606 | 0.444 | 1.365 | 0.172 | 1.833 | 0.791 | 4.615 |
| Non-Hispanic Black | 0.719 | 0.398 | 1.805 | 0.071 | 2.052 | 0.990 | 4.812 |
| Education Level |  |  |  |  |  |  |  |
| Below high school | Ref |  |  |  |  |  |  |
| High School Grad/GED or Equivalent | 0.161 | 0.327 | 0.494 | 0.622 | 1.175 | 0.618 | 2.239 |
| Some College or AA degree | -0.177 | 0.309 | -0.575 | 0.566 | 0.838 | 0.459 | 1.548 |
| College Graduate or above | -0.298 | 0.364 | -0.819 | 0.413 | 0.742 | 0.357 | 1.503 |
| Marital Status |  |  |  |  |  |  |  |
| Married/living with a partner | Ref |  |  |  |  |  |  |
| Never married | -0.713 | 0.334 | -2.134 | 0.0328* | 0.490 | 0.244 | 0.913 |
| Divorced | 0.645 | 0.304 | 2.124 | 0.0337* | 1.906 | 1.030 | 3.408 |
| Widowed/separated | -0.4673 | 0.542 | -0.863 | 0.388 | 0.627 | 0.184 | 1.622 |
| Family income-to-poverty ratio | -0.075 | 0.074 | -1.006 | 0.314 | 0.928 | 0.799 | 1.071 |
| *Examination data* |  |  |  |  |  |  |  |
| BMI (kg/m<sup>2</sup>) | 0.068 | 0.012 | 5.694 | 0.001* | 1.070 | 1.046 | 1.096 |
| *Laboratory data* |  |  |  |  |  |  |  |
| White blood cell count (1000 cells/uL) | 0.075 | 0.046 | 1.622 | 0.105 | 1.078 | 0.982 | 1.179 |
| Red blood cell count (million cells/uL) | 0.217 | 0.210 | 1.032 | 0.302 | 1.243 | 0.816 | 1.864 |
| Hemoglobin (g/dL) | 0.360 | 0.132 | 2.734 | 0.006* | 1.433 | 1.119 | 1.875 |
| Platelet count (1000 cells/uL) | 0.001 | 0.001 | 0.531 | 0.596 | 1.001 | 0.998 | 1.003 |
| Lymphocyte number (1000 cells/uL) | 0.115 | 0.154 | 0.745 | 0.456 | 1.122 | 0.819 | 1.501 |
| Monocyte number (1000 cells/uL) | 1.424 | 0.529 | 2.693 | 0.007* | 4.154 | 1.441 | 11.604 |
| Albumin (g/L) | 0.020 | 0.027 | 0.756 | 0.449 | 1.021 | 0.969 | 1.078 |
| Uric acid (umol/L) | 0.006 | 0.001 | 5.216 | 0.001* | 1.006 | 1.004 | 1.008 |
| Lactate dehydrogenase LDH (U/L) | 0.008 | 0.003 | 2.695 | 0.007* | 1.008 | 1.002 | 1.014 |
| Direct HDL-Cholesterol (mmol/L) | -0.832 | 0.311 | -2.677 | 0.007* | 0.435 | 0.232 | 0.785 |
| *Questionnaire data* |  |  |  |  |  |  |  |
| Asthma |  |  |  |  |  |  |  |
| Yes | Ref |  |  |  |  |  |  |
| No | 1.291 | 0.257 | 5.025 | 0.001* | 3.636 | 2.178 | 5.982 |
| Overweight |  |  |  |  |  |  |  |
| Yes | Ref |  |  |  |  |  |  |
| No | 1.905 | 0.269 | 7.084 | 0.001* | 6.721 | 4.038 | 1.652 |
| Arthritis |  |  |  |  |  |  |  |
| Yes | Ref |  |  |  |  |  |  |
| No | 1.652 | 0.248 | 6.677 | 0.001* | 5.219 | 3.204 | 8.473 |
| Congestive heart failure |  |  |  |  |  |  |  |
| Yes | Ref |  |  |  |  |  |  |
| No | 2.226 | 0.427 | 5.217 | 0.001* | 9.264 | 3.979 | 21.596 |
| Stroke |  |  |  |  |  |  |  |
| Yes | Ref |  |  |  |  |  |  |
| No | 1.709 | 0.498 | 3.429 | 0.001* | 5.521 | 1.981 | 14.442 |
| High blood pressure |  |  |  |  |  |  |  |
| Yes | Ref |  |  |  |  |  |  |
| No | 1.681 | 0.243 | 6.909 | 0.001* | 5.369 | 3.354 | 8.729 |
| Diabetes |  |  |  |  |  |  |  |
| Yes | Ref |  |  |  |  |  |  |
| No | 1.731 | 0.267 | 6.472 | 0.001* | 5.645 | 3.320 | 9.502 |
| Smoking |  |  |  |  |  |  |  |
| Yes | Ref |  |  |  |  |  |  |
| No | 0.593 | 0.236 | 2.511 | 0.012* | 1.809 | 1.133 | 2.867 |
| Urinary leakage |  |  |  |  |  |  |  |
| Yes | Ref |  |  |  |  |  |  |
| No | 0.309 | 0.241 | 1.281 | 0.200 | 1.362 | 0.841 | 2.172 |

**Table 3.** Weighted multivariate logistic regression analysis results of sleep disorder of research subjects in development cohort.

| Variables | Estimate | Std. Error | Z value | P-value | OR | CI (2.5%) | CI (97.5%) |
| --- | --- | --- | --- | --- | --- | --- | --- |
| Sex |  |  |  |  |  |  |  |
| Male | Ref |  |  |  |  |  |  |
| Female | -0.697 | 0.316 | -2.205 | 0.027 | 0.50 | 0.27 | 0.93 |
| Monocyte number (1000 cells/uL) | 1.637 | 0.631 | 2.595 | 0.009 | 5.14 | 1.49 | 17.7 |
| Asthma |  |  |  |  |  |  |  |
| Yes | Ref |  |  |  |  |  |  |
| No | 0.918 | 0.305 | 3.007 | 0.003 | 2.50 | 1.38 | 4.55 |
| Overweight |  |  |  |  |  |  |  |
| Yes | Ref |  |  |  |  |  |  |
| No | 1.295 | 0.355 | 3.647 | <0.001 | 3.65 | 1.82 | 7.32 |
| Arthritis |  |  |  |  |  |  |  |
| Yes | Ref |  |  |  |  |  |  |
| No | 0.795 | 0.304 | 2.616 | 0.009 | 2.22 | 1.22 | 4.02 |
| Congestive heart failure |  |  |  |  |  |  |  |
| Yes | Ref |  |  |  |  |  |  |
| No | 1.192 | 0.558 | 2.136 | 0.033 | 3.29 | 1.10 | 9.83 |
| Hypertension |  |  |  |  |  |  |  |
| Yes | Ref |  |  |  |  |  |  |
| No | 0.747 | 0.297 | 2.513 | 0.012 | 2.11 | 1.18 | 3.78 |

The construction of a predictive model was informed by the results of LASSO and logistic regression analyses, incorporating seven predictor variables (gender, monocyte count, asthma, overweight, arthritis, congestive heart failure, and hypertension). The model is represented in the form of a naming diagram (see Figure 3). A vertical line is then drawn from the top of the naming diagram to the corresponding variable, thus yielding the respective score. The scores for each variable are then summed to yield a total score, and the corresponding total risk score is placed at the bottom of the naming diagram. To illustrate, a 45-year-old male with a healthy body mass index, who is afflicted with arthritis and asthma but not hypertension or congestive heart failure, and has a monocyte count of 1, would have a total score of 140.5 points (gender 24 points, monocyte count 56 points, normal weight 0 points, arthritis 28 points, history of asthma 32.5 points, no hypertension 0 points, no congestive heart failure 0 points). The patient’s total score is 140.5, which corresponds to a probability of 0.3 to 0.4 in the record. The estimated probability of sleep disorders in this middle-aged and young adult population ranges from 30% to 40%.

**Figure 3.**
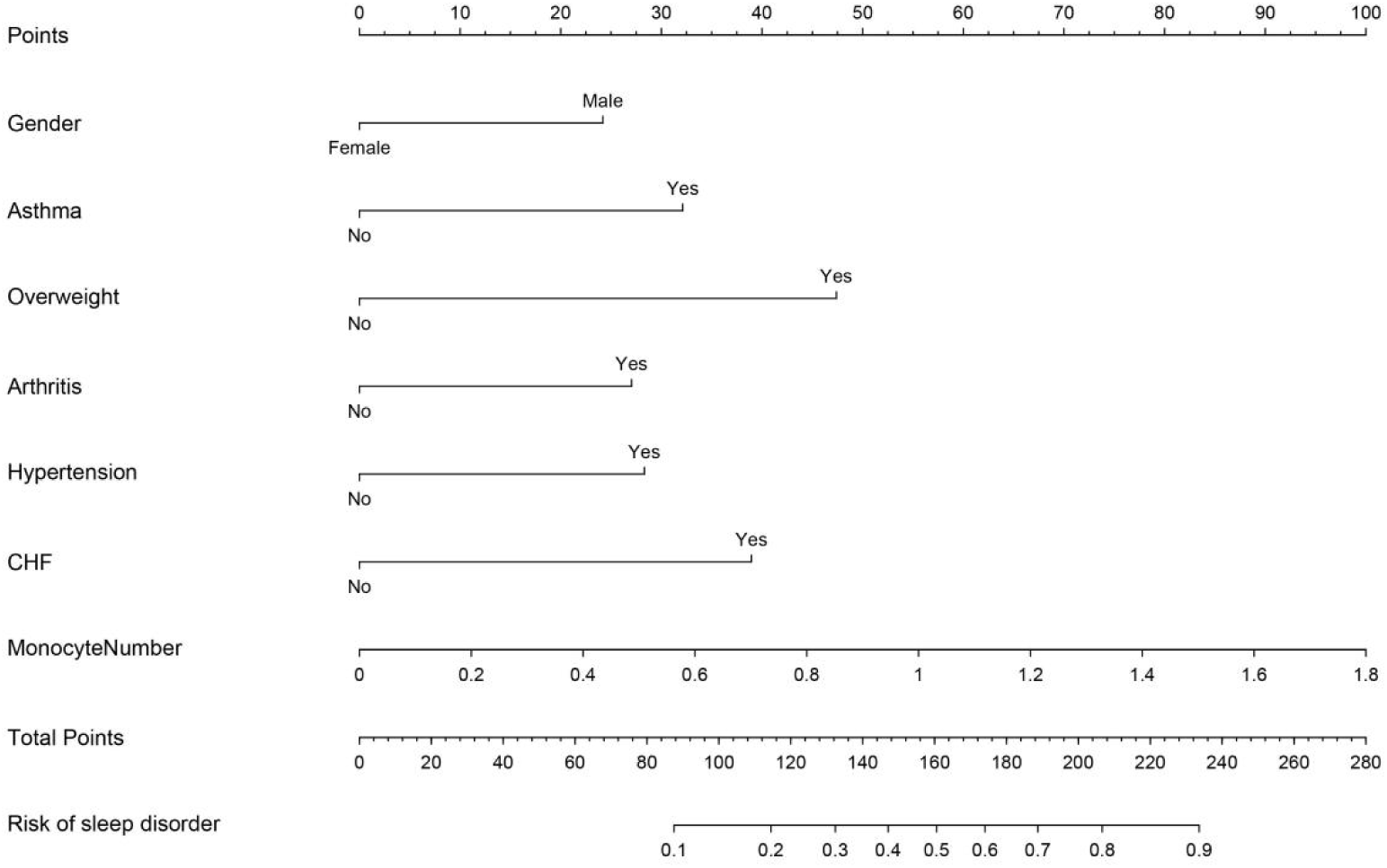
Nomogram model of the risk of sleep disorder in young and middle-aged patients.

### 3.3 Nomogram model evaluation

The validity and discriminatory power of this naming map were evaluated using multiple metrics. The AUC was utilized to evaluate the discriminatory capability of the scoring model. The AUC values for the scoring model on the development and validation cohorts were 0.817 and 0.791, respectively, suggesting that the model exhibited adequate discriminatory power (see Figure 4). The model’s calibration was evaluated through the utilisation of Brier scores and Spiegelhalter’s z-test. The Brier scores for the development and validation cohorts were 0.074 and 0.075, respectively, indicating that the model is well-calibrated. Spiegelhalter’s z-test demonstrated that the discrepancy between the probability of sleep disorders as predicted by the nomogram and the observed frequency of sleep disorders in patients was not statistically significant (*P* = 0.699, 0.718), thereby indicating that the model exhibits adequate predictive consistency (see Figure 5). The DCA was utilised to assess the clinical validity of the model (see Figure 6). The results of the DCA demonstrate that the predictive model yielded a net benefit for both the development and validation cohorts, thus indicating that the directory graph model offers superior net benefit and predictive accuracy.

**Figure 4.**
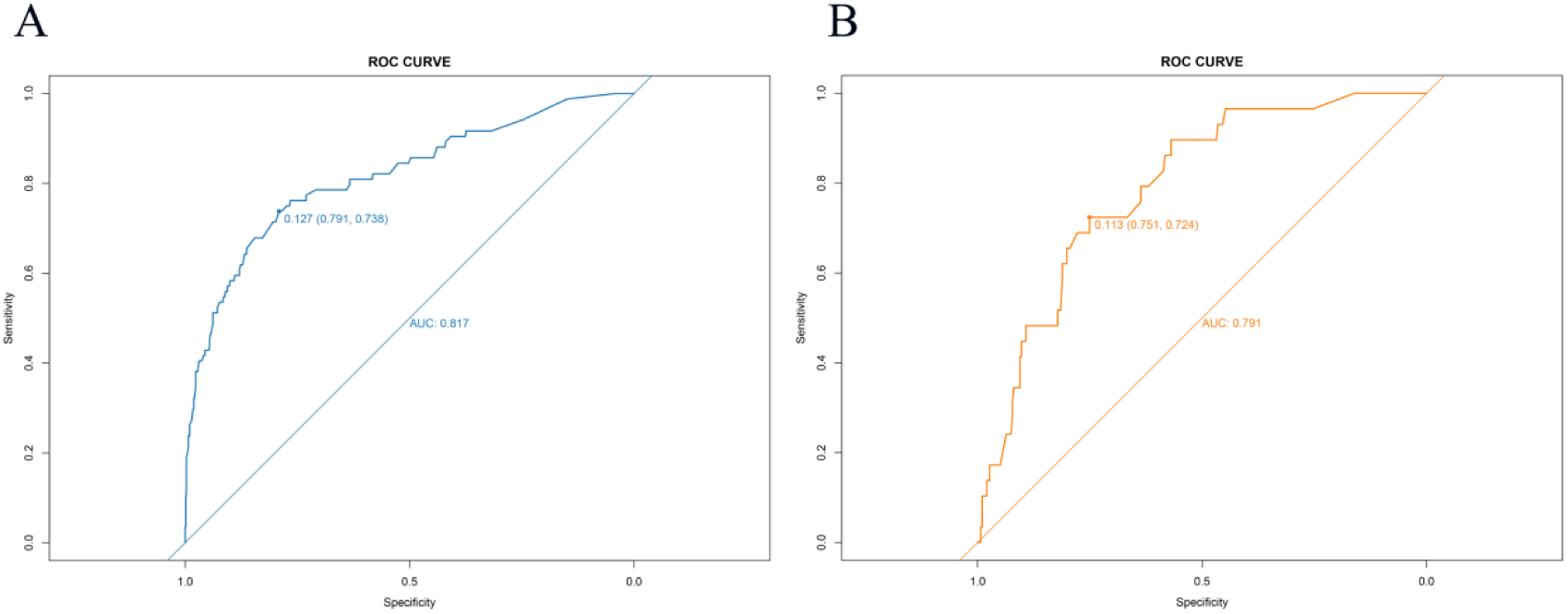
ROC curve of the nomogram model. Note: A. ROC curve of the nomogram model in development cohort; B. ROC curve of the nomogram model in validation cohort.

**Figure 5.**
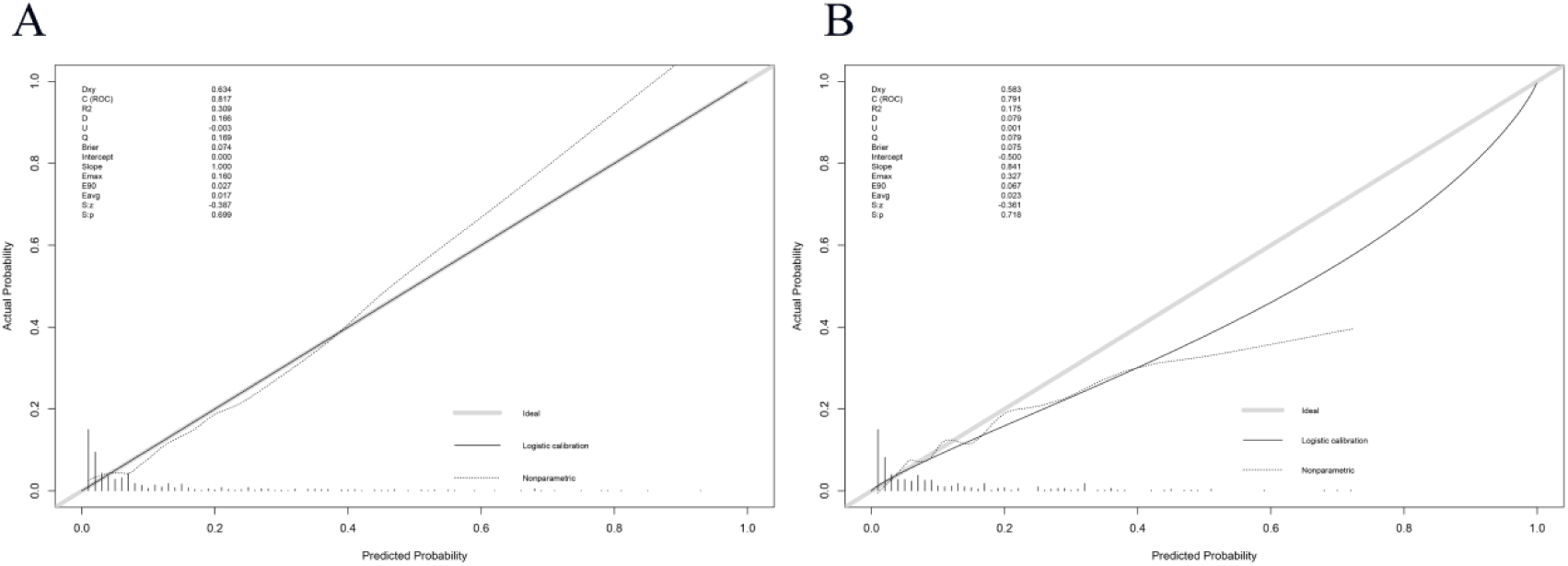
Calibration curve of nomogram model. Note: A. Calibration curve of the nomogram model in development cohort; B. Calibration curve of the nomogram model in validation cohort.

**Figure 6.**
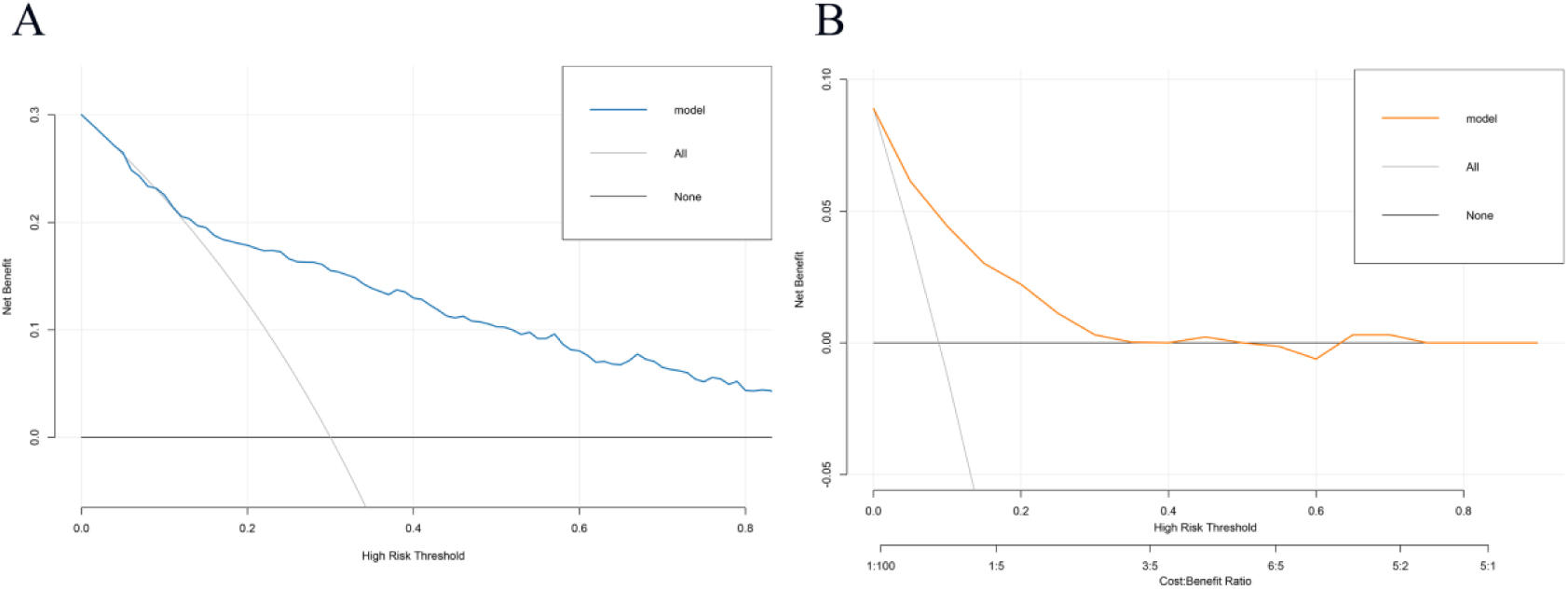
DCA curve of the nomogram model. Note: A. DCA curve of the nomogram model in development cohort; B. DCA curve of the nomogram model in validation cohort.

## 4 Discussion

Sleep disorders are not a single disease but rather a group of clinical conditions characterized by abnormalities in the quantity, quality, rhythm, or structure of sleep. These conditions often manifest as difficulty falling asleep, difficulty maintaining sleep, early awakening, non-restorative sleep, daytime sleepiness, and impaired daytime functioning [17,18]. In this study, sleep disorders were operationally defined based on self-reported sleep difficulties from the NHANES database. This definition more closely aligns with sleep difficulties or the risk of sleep disorders in epidemiological studies, rather than being entirely equivalent to specific sleep disorders diagnosed via polysomnography.

Sleep disorders have become a significant public health issue among young and middle-aged adults, with prevalence rates varying significantly depending on the study population, diagnostic criteria, and assessment tools. A plethora of studies have previously indicated that the prevalence of insomnia, defined by strict diagnostic criteria, is approximately 12.4%. However, estimates based on various sleep questionnaires have been found to range from 7.5% to 32.3% [19]. The Global Burden of Disease study suggests that the overall prevalence of insomnia among adults is approximately 16.2% [20]. McArdle et al. [21] found that among young adults with a mean age of 22.2 years, the prevalence of sleep disorders was 41.0% for women and 42.3% for men. Consequently, young and middle-aged adults are not a low-risk group for sleep disorders. A multitude of factors have been posited as potential contributors to an increased prevalence of sleep health issues, including academic and occupational stress, screen time, shift work, obesity, and the premature onset of chronic diseases. It has been demonstrated by preceding studies that an inadequate duration of sleep is associated with an elevated risk of cardiovascular disease, cognitive decline, metabolic abnormalities, depression, stroke, and all-cause mortality [22].

In the present study, sleep disorders were identified in 113 out of 1,089 middle-aged and young patients with anemia-representing a weighted prevalence rate of 10.40%. Whilst this result is lower than the overall prevalence of sleep problems reported in some studies of the general population, its clinical significance should not be overlooked, given that this study employed a specific operational definition and was limited to middle-aged and young patients with anemia.

The potential for a bidirectional relationship between anemia and sleep disorders is a salient consideration. Anemia can lead to a range of symptoms, including fatigue, weakness, decreased attention, reduced exercise tolerance, and impaired daytime functioning. These symptoms overlap significantly with manifestations of sleep disorders and may also amplify patients’ subjective perception of reduced sleep quality. Conversely, sleep disorders have been demonstrated to exacerbate fatigue, mood swings, cognitive decline, and impaired quality of life. The findings of the weighted multivariate logistic regression analysis in this study suggest that gender, monocyte count, asthma, overweight, arthritis, congestive heart failure, and hypertension are independent risk factors for sleep disorders in middle-aged and young patients with anemia.

The present study found that male patients suffering from anemia were more likely to have sleep disorders. As demonstrated in previous studies, males are especially prone to obstructive sleep apnea, short sleep duration and certain objective sleep disorders [23]. The relationship between monocyte count and sleep disorders is primarily associated with inflammation-related immune system pathways. The present study found that an elevated monocyte count was associated with an increased risk of sleep disorders. Al Rashed et al. found that sleep deprivation can affect monocyte subsets and inflammatory response, suggesting that monocytes may be the key cell bridge between sleep deprivation and systemic inflammation [22]. In obstructive sleep apnoea, intermittent hypoxia and sleep fragmentation have been demonstrated to induce oxidative stress, endothelial damage and monocyte activation. Polasky et al. discovered the redistribution of monocyte subsets in patients with obstructive sleep apnoea (OSA) [25]. Pries et al. also observed the changes of circulating CD14/CD16 monocyte subsets before and after OSA treatment [26]. It has been demonstrated that symptoms such as cough, wheezing, chest distress and airway hyper-responsiveness in patients with asthma have the capacity to directly destroy sleep continuity during nocturnal hours. Conversely, a state of sleep deprivation and a circadian rhythm disorder have been shown to be capable of aggravating airway inflammation and immune response. Liu et al. found that sleep disorders were associated with an increased prevalence and risk of asthma [27]. Kavanagh et al. also highlighted that inadequate sleep quality and sleep disorders are prevalent among patients with asthma and interact with poor asthma control [28]. Overweight or obesity is considered to be one of the most significant intervention risk factors for the development of sleep disorders. Wang et al. found that BMI level is associated with an increased risk of sleep disorders, as evidenced by data from the NHANES survey from 2005 to 2018. Obese individuals were found to have a higher probability of developing sleep disorders [29]. Parmelee and other studies have demonstrated that sleep disorders in patients with osteoarthritis are significantly associated with pain and depression, and can predict subsequent depression and exacerbation of dysfunction [30], suggesting that the relationship between arthritis and sleep disorders is primarily facilitated by pain, inflammation and psychological burden. The relationship between congestive heart failure and sleep disorders is more complicated. Laugsand et al. demonstrated that insomnia symptoms were associated with an elevated risk of new-onset heart failure [31]. Redeker et al. noted that insomnia is prevalent among patients with stable heart failure and impacts daytime function [32]. Relevant reviews also assert that insomnia is highly prevalent among patients with chronic heart failure and is closely related to fatigue and a decline in quality of life [33]. The relationship between hypertension and sleep disorders is relatively stable, especially in cases of short sleep, insomnia and sleep apnoea. The meta-analysis of prospective cohort by Li et al. demonstrated that sleep disorders were associated with an elevated risk of hypertension [34], a finding that is in accordance with the results of the present study.

In comparison with assessments based on single risk factors, risk charts have the capacity to integrate demographic characteristics, laboratory indicators, comorbidities, and symptom variables, thereby transforming complex models into intuitive, actionable, and personalized risk assessment tools [35]. In recent years, there has been an increasing use of risk charts for predicting the risk of sleep disorders [36].

However, to date, no studies have reported risk prediction models for sleep disorders in young and middle-aged patients with anemia. In order to address this gap in the literature, a nomogram was constructed in order to predict the probability of sleep disorders in middle-aged and young patients with anemia using seven predictors that were selected via LASSO regression analysis. The nomogram exhibited satisfactory discriminatory capability, with an area under the ROC curve of 0.817 in the development cohort and 0.791 in the validation cohort. The values thus obtained are indicative of the model’s capacity to effectively differentiate between patients with and without sleep disorders. In addition, the model’s Brier score was determined to be 0.074 in the development cohort and 0.075 in the validation cohort, suggesting a high degree of calibration. Furthermore, the DCA demonstrated clinical net benefit within a certain threshold probability range, suggesting that the model possesses good discriminatory power, calibration, and clinical applicability.

The present study is subject to several limitations. Firstly, NHANES is a cross-sectional survey, which makes it difficult to infer causal relationships [34]. Secondly, some variables rely on self-reporting, which may introduce recall bias and information bias [37]. In addition, the model has solely undergone internal validation [35], and the generalisability of the results necessitates further validation in external cohorts. In order to enhance the model’s stability, interpretability, and clinical utility, prospective multicentre studies are required.

## 5 Conclusion

The present study employed both LASSO regression and multivariate logistic regression in order to identify potential predictive factors. In the final analysis, seven predictive variables-including gender, monocyte count, asthma, overweight, arthritis, congestive heart failure, and hypertension-were incorporated into the nomogram. The predictive model demonstrates good predictive performance and clinical applicability, based on independent risk factors for sleep disorders across participants. The primary function of the tool is to assist clinicians in conducting early risk stratification and developing personalised intervention strategies that target modifiable risk factors.

## Ethics approval and consent to participate

This study adhered to the principles outlined in the Declaration of Helsinki. The NHANES data used in this research were approved by the Institutional Review Board (IRB) of the NCHS. All participants gave written informed consent to participate in this study.

## Acknowledgements

The authors would like to express their gratitude to the NHANES participants and staff for their contributions to the data and data collection.

## Authors’ contributions

Y.L. designed the study and wrote the manuscript. C.Z. drafted and revised the manuscript. All authors reviewed and approved the final version of the manuscript.

## Data availability

The datasets analyzed during the current study are available in the NHANES repository, https://www.cdc.gov/nchs/nhanes/.

## Consent for publication

Not applicable.

## Clinical trial number

Not applicable.

